# The Hypoxic Burden of Sleep Apnea Predicts Cardiovascular Outcomes in Patients with Heart Failure

**DOI:** 10.64898/2026.09.29.26364138

**Authors:** Minori Masaki, Saki Tsumoto, Nanako Shiroshita, Fusae Kawana, Sayaki Ishiwata, Hiroyuki Kitagawa, Masashi Yanagisawa, Takatoshi Kasai

**Affiliations:** International Institute for Integrative Sleep Medicine (WPI-IIIS), Tsukuba Institute for Advanced Research, University of Tsukuba, Ibaraki, Japan; PhD program in Humanics, University of Tsukuba, Ibaraki, Japan; Department of Cardiovascular Respiratory Sleep Medicine, Juntendo University Graduate School of Medicine, Tokyo, Japan; Yumino Heart Clinic, Tokyo, Japan; Department of Cardiovascular Biology and Medicine, Juntendo University Graduate School of Medicine, Tokyo, Japan; S’UIMIN Inc., Tokyo, Japan

**Keywords:** apnea-hypopnea index, hypoxic burden, acute heart failure, polysomnography, positive airway pressure, prognosis

## Abstract

**Background:** Sleep-disordered breathing (SDB) is an established adverse prognostic factor in heart failure (HF). Sleep apnea-specific hypoxic burden (SASHB), which reflects the duration and depth of respiratory event–related oxygen desaturation, has shown stronger prognostic value than the apnea-hypopnea index (AHI) in general populations and patients with coronary artery disease. However, its clinical utility in patients hospitalized due to acute HF remains unclear.

**Objectives:** This study evaluated the prognostic value of SASHB in patients hospitalized for acute HF and compared its performance with AHI for risk stratification and identification of patients who may benefit from positive airway pressure (PAP) therapy.

**Methods:** This post hoc analysis included 230 patients from a prospective single-center observational cohort of adults hospitalized for acute HF. The primary analysis included 178 patients who did not receive PAP treatment. Associations of SASHB and AHI with the composite endpoint of cardiovascular death or rehospitalization for worsening HF were evaluated using Cox proportional hazards models. Model improvement after adding SASHB or AHI to a model including established clinical risk factors was assessed using changes in the C index and Brier score, reflecting changes in model discrimination and prediction error, respectively. In the full cohort, the interaction between PAP treatment and SDB metrics was evaluated using the g-formula.

**Results:** During a median follow-up of 3.7 years, 58 of 178 patients (32.6%) reached the composite endpoint. Both SASHB and AHI were independently associated with the composite outcome after multivariable adjustment. However, only SASHB consistently predicted outcomes across analyses and improved model performance beyond established prognostic factors (bootstrap ΔC index, 0.020 [95% CI: 0.008–0.031]; bootstrap ΔBrier score, −0.007 [95% CI: −0.014 to −0.001]), whereas AHI did not. In the analysis including PAP-treated patients, both SASHB and AHI showed interaction with PAP treatment.

**Conclusions:** In patients hospitalized for acute HF, SASHB showed a consistent independent association with adverse clinical outcomes and provided additional prognostic information beyond AHI. SASHB may improve risk stratification and may help identify patients who are more likely to benefit from PAP therapy.

## Introduction

Heart failure (HF) remains associated with high mortality and rehospitalization rates despite advances in treatment (1,2). Identifying patients with HF at high risk and mitigating modifiable risk factors are therefore critical for improving clinical outcomes. Sleep-disordered breathing (SDB) is an established adverse prognostic factor in HF, either acute or chronic HF (3–5), and may represent a favorable therapeutic target because its associated risk may be reduced by positive airway pressure (PAP) treatment (3,6,7), whereas the efficacy of PAP treatment for moderate to severe SDB on prognosis in patients with acute or chronic HF has not yet been proven in randomized controlled trials (8–11). Apnea-hypopnea index (AHI) has traditionally been used as the standard metric for assessing SDB severity and has been associated with increased mortality risk (7,12,13). However, AHI provides only the frequency of respiratory events per hour of sleep and does not account for the duration or depth of ventilatory disturbance or blood gas changes; it may capture only one aspect of the physiological burden of SDB. Sleep apnea-specific hypoxic burden (SASHB) was developed to quantify the duration and depth of oxygen desaturation associated with respiratory events (14). Prior studies evaluating SDB metrics in general populations (14,15) and in patients with coronary artery disease (16) have suggested that SASHB may provide stronger prognostic information than AHI. However, the clinical utility of SASHB in patients with either acute or chronic HF and the effects of SASHB for determining patients who may have mortality and morbidity benefits with PAP treatment remain to be elucidated. Therefore, we investigated whether SASHB is associated with adverse clinical outcomes in patients hospitalized due to HF and whether it may be useful as a metric for the indication of PAP treatment for SDB in them.

## Method

### Study Design

This study was a post hoc analysis of a previously conducted single-center observational cohort that examined the association between SDB or PAP treatment for SDB and the risk of cardiovascular death or rehospitalization for HF in patients hospitalized for acute HF (7). In the present analysis, we evaluated the prognostic value of SASHB and AHI and their potential relevance to PAP treatment.

The Research Ethics Committee, Faculty of Medicine, Juntendo University approved the study protocol for this post hoc analysis study protocol (E24-0457-H01), and the study complied with the Declaration of Helsinki. Written informed consent was obtained from all patients. Since all data had already been obtained, the requirement to obtain informed consent was waived by the Research Ethics Committee, Faculty of Medicine, Juntendo University, using opt-out methods.

### Study populations

Trial details have been summarized previously (7). In brief, the present study used patient data from a previously conducted observational study of patients hospitalized for acute HF at our institution (Juntendo University Hospital, Tokyo, Japan) between May 2012 and April 2018. Patients with acute HF who met the modified Framingham criteria, which included only variables assessed at admission, were eligible for the original study (17,18). After initial improvement of acute signs of HF, overnight polysomnography was performed to assess eligibility according to predefined inclusion and exclusion criteria.

The original study included men and women aged ≥ 20 years with left ventricular ejection fraction (LVEF) < 50% on echocardiography. Patients were excluded if they required oxygen therapy, had acute coronary syndrome or cardiac surgery within the previous 4 weeks, had end-stage renal disease requiring dialysis, cerebrovascular disease with neurological deficits, life-threatening malignancy, apparent obstructive lung disease, or known SDB. Of the 241 patients included in the original study, 11 were excluded because of inadequate oxyhemoglobin saturation by pulse oximeter (SpO_2_) measurement or data corruption; therefore, 230 patients were included in the present analysis.

### Sleep Studies and PAP Treatment

This study used polysomnography data obtained in the original cohort (7). All patients underwent overnight polysomnography using Alice PDX (Philips Respironics, Murrysville, PA, USA) during the first hospitalization for acute HF after initial improvement of acute signs and symptoms. Electrocardiography, electroencephalography, electrooculography, and electromyography were performed. Thoracoabdominal motion was monitored using respiratory inductance plethysmography; airflow was measured using an oronasal thermal airflow sensor and nasal pressure cannula; and SpO_2_ was monitored using pulse oximetry. Respiratory events were defined and scored according to the American Academy of Sleep Medicine manual version 2.2. In this study, SDB was defined as an AHI ≥ 15 events/h. Apneas were classified as obstructive sleep apnea (OSA) or central sleep apnea (CSA) according to the presence or absence of thoracoabdominal motion, respectively. Hypopneas were classified as central if none of the following criteria were met: snoring during the event, increased inspiratory flattening of the nasal pressure compared to baseline breathing, and associated thoracoabdominal paradox during but not before the event (19).

PAP treatment, including continuous PAP or adaptive servo ventilation, was initiated according to the decision of the patient or attending physicians.

### Clinical Data Collection

Baseline data were collected prospectively at the time of the sleep study, and medical history was obtained by chart review. New York Heart Association (NYHA) functional class was assessed at the time of the sleep study. Renal function was expressed as estimated glomerular filtration rate (eGFR), calculated from baseline serum creatinine levels using the Modification of Diet in Renal Disease Study equation with the Japanese coefficient (20). Echocardiography was performed at the time of the sleep study, and LVEF was calculated using the modified Simpson method (21).

### Derivation and Categorization of SASHB, AHI and T90

SASHB was calculated according to published methods (14). Briefly, respiratory events were manually classified as apnea or hypopnea and as obstructive, central, or mixed. Oxygen desaturation curves associated with all individually identified events were aligned and averaged across the entire night to generate a mean desaturation curve. In the mean desaturation curve, the interval from the onset of desaturation to recovery was defined as the search window, corresponding to the desaturation duration. This search window was then applied to each event to calculate the oxygen desaturation area. SASHB was calculated as the sum of these event-specific desaturation areas divided by total sleep time. This procedure could be applied to both obstructive and central events without modification, allowing SASHB to be calculated for all respiratory events combined and separately by event class and type (Supplemental Figure 1A-D).

Obstructive and central AHI were also computed separately. In addition, patients with SDB were classified into an obstructive-dominant group (i.e., ≥ 50% obstructive events) and a central-dominant group (i.e., ≥ 50% central events) (22). Time with oxygen saturation < 90% (T90) was defined as the total time, in minutes, during the recording period with oxygen saturation < 90% (23–25).

Consistent with the conceptual approximation SASHB ≈ AHI × (desaturation depth × desaturation duration)/2, a power-law relationship was observed between SASHB, an area-based metric, and AHI, a frequency-based metric (SASHB = 0.836 × AHI^1.31; Supplemental Figure 2A). Therefore, we use the square root-transformed SASHB (sqrt HB) (14), which compresses the scale of SASHB, for comparison with AHI. Considering the threshold of AHI ≥ 30 events/h for severe SDB and the median values in the present study population (AHI, 30.7 events/h; sqrt HB, 7.9), the cutoff values were set at 8 for sqrt HB and 30 for AHI. For T90, a cutoff of 5 min was selected based on the median value in the present study (5.09 min) and previous reports (23).

### Follow-up and outcomes

Outcome data were re-reviewed in February 2026 by examining medical records at our hospital in a blinded manner. We followed all patients from the date of the sleep study during the initial hospitalization, and administrative censoring was applied at 5 years because deaths from causes other than HF were expected to increase over longer follow-up; therefore, the latest follow-up date used in the present analysis was April 2023. The endpoint of interest was the composite of cardiovascular death or rehospitalization for worsening HF.

### Statistical methods

Continuous variables are presented as mean ± standard deviation (SD), and categorical variables are presented as numbers and percentages. Baseline characteristics were compared between groups using the chi-squared test for categorical variables and the Mann–Whitney U test for continuous variables. For each continuous variable, the effect size of the between-group difference was expressed as Cohen’s *d*. Event-free survival curves were generated using the Kaplan–Meier method and compared between groups using the log-rank test. Both univariable and multivariable Cox models in this study fulfilled the proportional hazards assumption. Associations between continuous variables were evaluated using simple linear regression after logarithmic transformation, as appropriate.

The primary analysis included patients who did not receive PAP treatment to avoid the potential influence of PAP treatment on outcomes. Multivariable Cox proportional hazards models included baseline factors considered clinically relevant. In the primary analysis, because of the number of incident events, the number of variables in the multivariable Cox models was limited to avoid overfitting. The baseline model included age, New York Heart Association (NYHA) functional class treated as a continuous variable, hemoglobin, log B-type natriuretic peptide (BNP), and eGFR (26,27). Models adding either sqrt HB or AHI to the baseline model were then compared with the baseline model. Both metrics were analyzed continuously and categorically using prespecified cutoffs; Hazard ratios (HR) for continuous analyses were expressed per 1-SD increase.

Model improvement after adding SASHB or AHI to a model including established prognostic factors was assessed using the change in the C index and the Brier score at 5 years (28,29). The Brier score was calculated using inverse probability of censoring weighting (30). To assess the stability of the incremental predictive performance, bootstrap resampling was performed with 500 iterations (31). In each iteration, the Cox models were refitted in a bootstrap sample and evaluated in the original analytic dataset. The bootstrap distribution of the difference in performance between the extended and baseline models was used to derive percentile-based 95% confidence intervals (CIs).

As a secondary analysis, the interaction between PAP treatment and SASHB or AHI was evaluated using the g-formula (32) in the full analytic dataset of 230 patients, including those treated with PAP. Because of the limited number of composite outcome events occurred among patients treated with PAP, the number of covariates was restricted to NYHA functional class and hemoglobin. Separate Cox proportional hazards models were fitted for SASHB and AHI, each including PAP treatment, the corresponding metric categorized using a cutoff of 8 for sqrt HB or 30 events/h for AHI, their interaction term, and the adjustment covariates. Using these models, the g-formula was applied by predicting each patient’s five-year risks, which were then standardized under hypothetical PAP treatment and no PAP treatment and averaged over the observed covariate distribution within the lower and higher SASHB or AHI categories. The absolute risk reduction associated with PAP treatment was calculated as the difference in standardized 5-year risk between the no-PAP and PAP scenarios within each category. As sensitivity analyses, all primary and secondary analyses were repeated using T90, evaluated both continuously and dichotomized at 5 min, the median value in this study.

All statistical analyses were performed using Python version 3.9.19 (Python Software Foundation, Wilmington, Delaware, USA). Survival analyses were conducted using the lifelines package 0.27.8 (33), statistical analyses using SciPy 1.7.0 (34), and data visualization using Seaborn 0.13.0 (35).

### Data-sharing Statement

The data underlying this study are not available for sharing because participant consent for data sharing was not obtained at the time the original study was initiated.

## Results

### Patient Population

Among patients included in a previously established database from a study evaluating SDB at admission and the association between PAP treatment and clinical outcomes in patients with HF (7), 230 patients were included in the analytic dataset after excluding those with partially inadequate SpO_2_ data. Of these, 178 patients who did not receive PAP treatment were included in the primary analysis, and all 230 patients were included in the secondary analysis evaluating the interaction between PAP treatment and SASHB or AHI (Figure 1).

**Figure 1.**
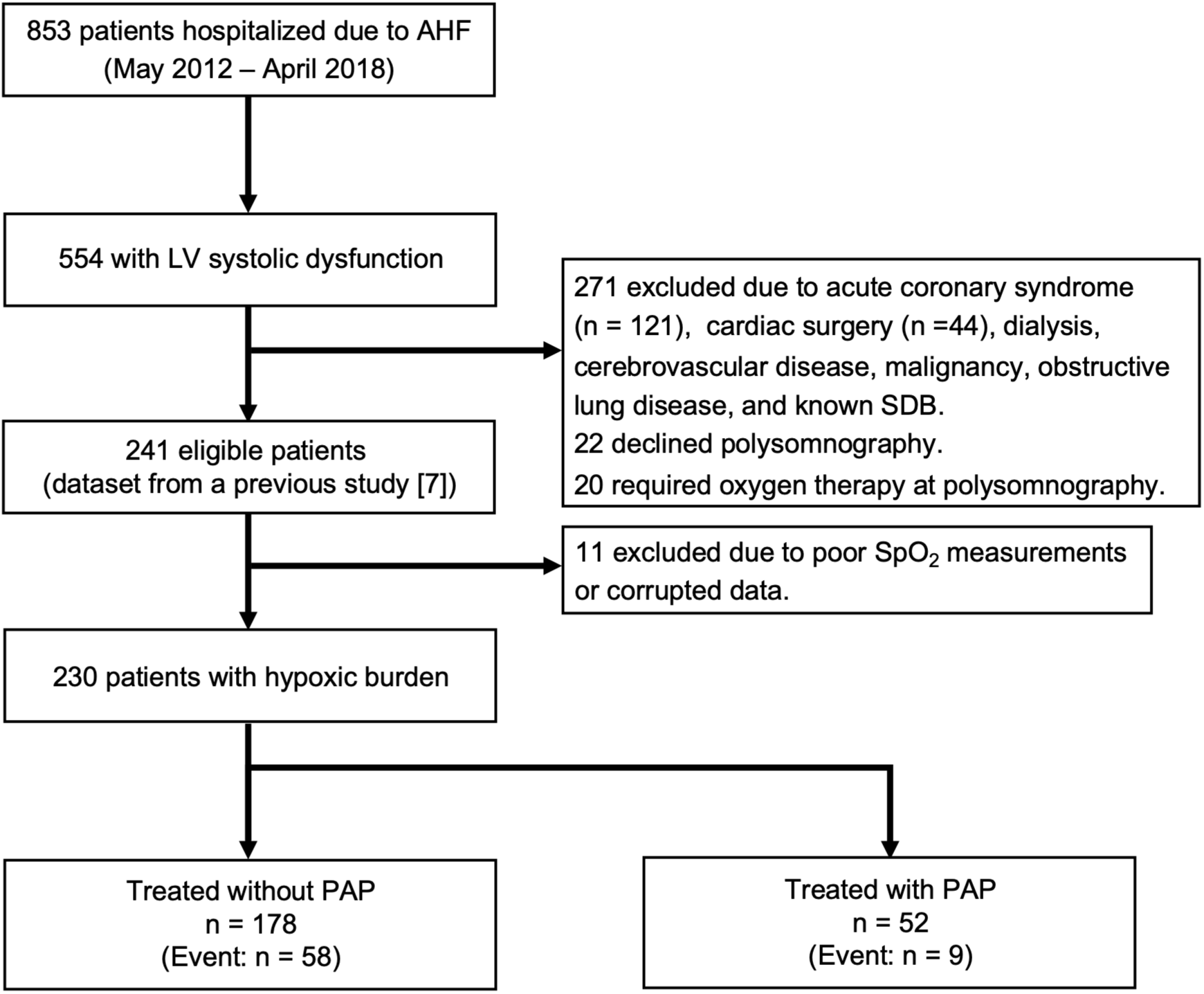
Flow diagram of the study population. From May 2012 to April 2018, 853 patients with acute HF were hospitalized at Juntendo University Hospital. Among 554 patients with LVEF < 50%, 271 were excluded according to prespecified criteria, and an additional 42 patients were excluded before or at polysomnography, leaving 241 patients in the dataset from the previous study (7). Of these, 11 were excluded because of inadequate SpO_2_ data. Thus, 230 patients were included in the present study: 178 without PAP treatment and 52 with PAP treatment. HF = heart failure; LVEF = left ventricular ejection fraction; PAP = positive airway pressure; SDB = sleep-disordered breathing; SpO_2_ = oxyhemoglobin saturation.

Table 1 summarizes the baseline characteristics of the 178 patients. The mean age was 63.9 ± 13.1 years, 127 patients were male (71.3%), and the mean body mass index (BMI) was 23.5 ± 4.7 kg/m^2^. Of the 178 patients, 123 (69.1%) had SDB. Among these patients, 70 (56.9%) were classified into the central-dominant group and 53 (43.1%) into the obstructive-dominant group.

**Table 1.** Clinical Characteristics by Endpoint Event Status. Values are mean ± SD or n (%). Clinical characteristics were compared between patients with and without endpoint events of cardiovascular death or rehospitalization for worsening HF, using the chi-squared test for categorical variables and the Mann–Whitney U test for continuous variables. ACE-I = angiotensin-converting enzyme inhibitor; AHI = apnea-hypopnea index; ARB = angiotensin II receptor blocker; BMI = body mass index; BNP = B-type natriuretic peptide; BP = blood pressure; CRT = cardiac resynchronization therapy; eGFR = estimated glomerular filtration rate; HF = heart failure; ICD = implantable cardioverter defibrillator; LVEF = left ventricular ejection fraction; NYHA = New York Heart Association; ODI = oxygen desaturation index; REM = rapid eye movement; SASHB = sleep apnea-specific hypoxic burden; SD = standard deviation; sqrt HB = square root-transformed SASHB; T90 = time with oxygen saturation < 90%; TRT = total recording time; TST = total sleep time.

| Variable | Total (n = 178) | Event free (n = 120) | Incident case (n =58) | p value | Cohen's d |
| --- | --- | --- | --- | --- | --- |
| Patient characteristics |  |  |  |  |  |
| Age, year | 63.9 ± 13.1 | 62.7 ± 13.2 | 66.4 ± 12.7 | 0.078 | 0.282 |
| Age ≥ 60, n(%) | 111 (62.36) | 69 (57.5) | 42 (72.41) | 0.054 |  |
| Male sex, n(%) | 127 (71.35) | 88 (73.33) | 39 (67.24) | 0.399 |  |
| BMI kg/m<sup>2</sup> | 23.5 ± 4.7 | 23.2 ± 4.4 | 24.3 ± 5.1 | 0.288 | 0.233 |
| BMI ≥ 25 kg/m<sup>2</sup>, n(%) | 51 (28.65) | 31 (25.83) | 20 (34.48) | 0.232 |  |
| History of HF, n (%) | 87 (48.88) | 59 (49.17) | 28 (48.28) | 0.911 |  |
| NYHA class at polysomnography | 2.4 ± 0.6 | 2.3 ± 0.5 | 2.5 ± 0.7 | **0.014** | 0.389 |
| NYHA III/IV, n (%) | 55 (30.9) | 30 (25.0) | 25 (43.1) | **0.014** |  |
| Systolic BP, mmHg | 105.3 ± 16.2 | 104.4 ± 15.4 | 107.1 ± 17.8 | 0.502 | 0.166 |
| Diastolic BP, mmHg | 59.8 ± 9.9 | 59.7 ± 10.1 | 60.1 ± 9.6 | 0.637 | 0.038 |
| Heart rate, /min | 71.1 ± 12.0 | 70.5 ± 11.7 | 72.3 ± 12.6 | 0.442 | 0.150 |
| LVEF, % | 34.4 ± 10.6 | 34.9 ± 11.1 | 33.6 ± 9.4 | 0.565 | -0.122 |
| ICD, n (%) | 13 (7.3) | 6 (5.0) | 7 (12.07) | 0.089 |  |
| CRT, n (%) | 14 (7.87) | 8 (6.67) | 6 (10.34) | 0.393 |  |
| Comorbidities |  |  |  |  |  |
| Atrial brillation, n (%) | 69 (38.76) | 42 (35.0) | 27 (46.55) | 0.138 |  |
| Diabetes, n (%) | 50 (28.09) | 31 (25.83) | 19 (32.76) | 0.335 |  |
| Laboratory variables |  |  |  |  |  |
| Haemoglobin, g/L | 13.5 ± 2.5 | 14.0 ± 2.5 | 12.4 ± 2.0 | **< 0.001** | -0.670 |
| Sodium, mmol/l | 139.5 ± 3.3 | 139.6 ± 3.6 | 139.3 ± 2.8 | 0.347 | -0.092 |
| Potassium, mmol/l | 4.6 ± 2.8 | 4.7 ± 3.4 | 4.4 ± 0.5 | 0.499 | -0.095 |
| BNP, pg/ml | 406.3 ± 382.8 | 338.8 ± 328.3 | 546.1 ± 447.9 | **< 0.001** | 0.558 |
| log BNP | 5.6 ± 1.0 | 5.4 ± 1.0 | 6.0 ± 0.9 | **< 0.001** | 0.579 |
| eGFR, ml/min/1.73 m<sup>2</sup> | 44.4 ± 19.5 | 47.9 ± 19.0 | 37.2 ± 18.8 | **< 0.001** | -0.567 |
| eGFR < 60, n (%) | 139 (78.09) | 89 (74.17) | 50 (86.21) | 0.069 |  |
| Sleep study findings |  |  |  |  |  |
| Total recording time, min | 623.0 ± 63.7 | 626.5 ± 70.6 | 615.8 ± 46.1 | 0.377 | -0.168 |
| Total sleep time, min | 355.3 ± 84.1 | 367.3 ± 79.0 | 330.6 ± 89.6 | **0.009** | -0.444 |
| Sleep efficiency, % of TST | 57.3 ± 13.3 | 58.9 ± 12.4 | 54.0 ± 14.7 | **0.042** | -0.372 |
| Wake after sleep offset, min | 147.7 ± 80.6 | 135.3 ± 71.6 | 173.2 ± 92.1 | **0.015** | 0.480 |
| Slow wave sleep, min | 25.6 ± 26.2 | 29.4 ± 28.3 | 17.7 ± 19.0 | **0.006** | -0.456 |
| REM sleep, min | 55.4 ± 30.5 | 58.9 ± 30.7 | 48.0 ± 29.2 | **0.017** | -0.361 |
| AHI, /h of sleep | 29.8 ± 19.3 | 28.0 ± 19.3 | 33.4 ± 19.0 | 0.080 | 0.280 |
| HB, %min/h of sleep | 83.2 ± 81.8 | 75.9 ± 78.4 | 98.5 ± 87.1 | **0.044** | 0.279 |
| sqrt HB | 8.1 ± 4.3 | 7.7 ± 4.2 | 8.9 ± 4.3 | **0.044** | 0.307 |
| 3% ODI, /h of sleep | 22.7 ± 17.9 | 20.9 ± 17.6 | 26.4 ± 18.2 | 0.075 | 0.305 |
| T90, min of TRT | 28.7 ± 64.3 | 26.0 ± 66.2 | 34.3 ± 60.5 | 0.075 | 0.129 |
| Obstructive-dominant, n (%) | 53 (29.8) | 33 (27.5) | 20 (34.5) | 0.340 |  |
| Medication at discharge |  |  |  |  |  |
| ACE-Is/ARBs | 142 (79.78) | 95 (79.17) | 47 (81.03) | 0.771 |  |
| Beta blocker | 166 (93.26) | 110 (91.67) | 56 (96.55) | 0.223 |  |
| Aldosterone antagonist | 111 (62.36) | 76 (63.33) | 35 (60.34) | 0.700 |  |
| Loop diuretics | 145 (81.46) | 93 (77.5) | 52 (89.66) | 0.050 |  |

Patients with central-dominant SDB had a significantly longer desaturation duration than those with obstructive-dominant SDB (Supplemental Figure 1F), and desaturation duration increased as the central event ratio increased (β = 0.186 [95% CI: 0.118–0.253]; *p* < 0.001) (Supplemental Figure 1G). Consistent with the longer desaturation duration, the mean area per event was also significantly greater in patients with central-dominant SDB (Supplemental Figure 1F).

During follow-up, 58 patients (32.6%) reached the composite endpoint, including 4 cardiovascular deaths and 54 rehospitalizations for worsening heart failure. The median follow-up duration was 3.7 years (95% CI: 2.2–4.7 years). Patients who experienced endpoint events had higher SASHB values than those who did not, whereas the AHI did not differ between the groups (Table 1).

The characteristics of patients stratified by sqrt HB ≥ 8 and AHI ≥ 30 events/h are shown in Supplemental Table 1. Patients in the high-sqrt HB and high-AHI groups had higher BMI and a higher prevalence of diabetes. The high-sqrt HB group had higher BNP levels, whereas the high-AHI group had worse NYHA functional class and lower eGFR.

### Association of sqrt HB and AHI With the Composite Outcome

Higher sqrt HB and AHI values were associated with an increased risk of cardiovascular death or rehospitalization for heart failure. Kaplan–Meier curves showed a higher cumulative incidence of the composite endpoint in patients with sqrt HB ≥ 8 and those with AHI ≥ 30 events/h (Figure 2). The estimated 5-year probabilities of the composite outcome were 27.5% and 49.6% in the low- and high-sqrt HB groups, respectively, and 28.6% and 47.0% in the low- and high-AHI groups, respectively.

**Figure 2.**
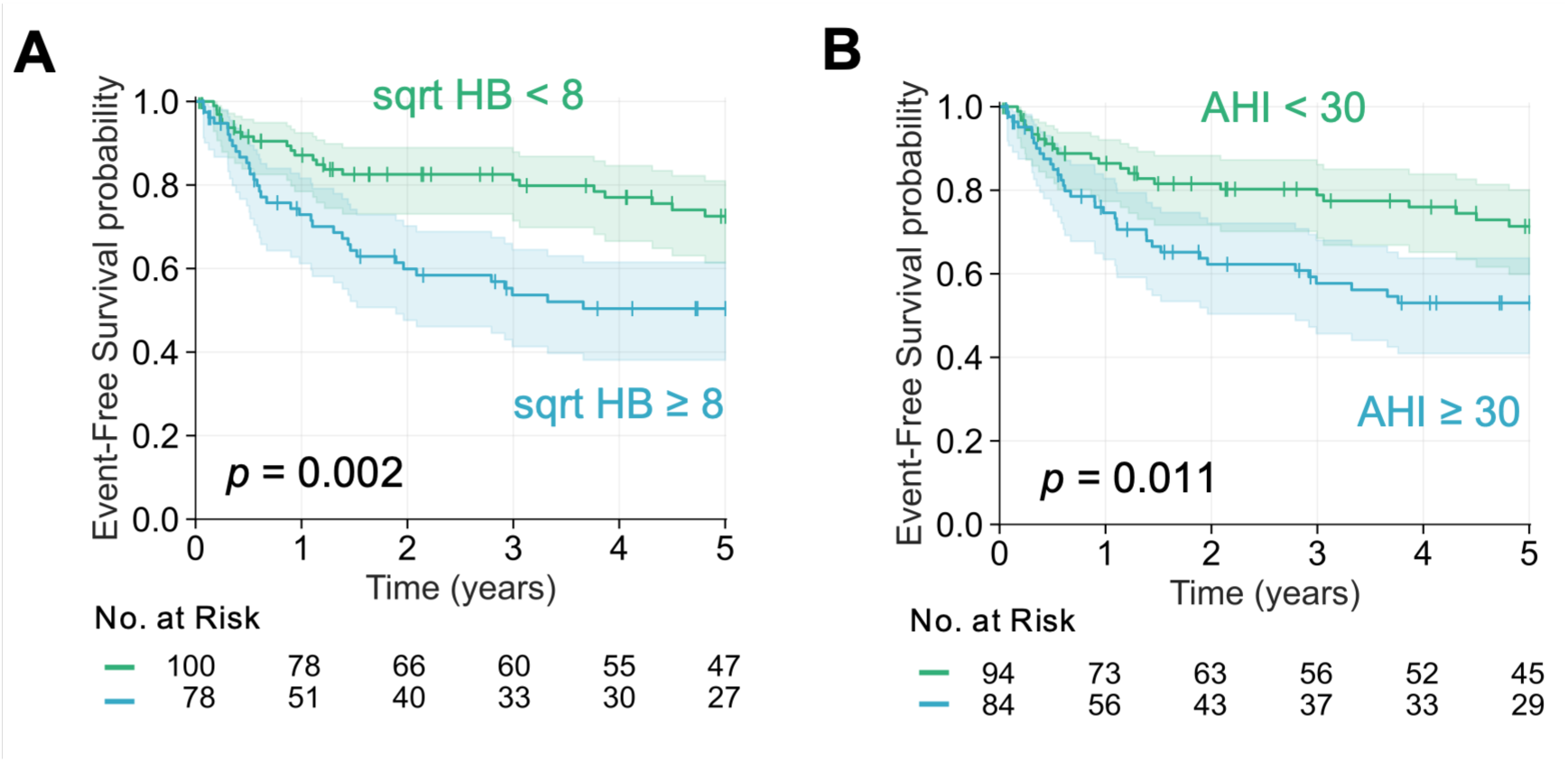
Kaplan–Meier event-free survival curves by SASHB and AHI. Kaplan–Meier event-free survival curves are shown according to the cutoffs of (A) sqrt HB ≥ 8 and (B) AHI ≥ 30 events/h. Cumulative event-free survival for the composite of cardiovascular death or rehospitalization for worsening HF was lower in patients with sqrt HB ≥ 8 or AHI ≥ 30 events/h than in their respective lower-value groups, and between-group differences were assessed using the log-rank test. AHI = apnea-hypopnea index; HF = heart failure; sqrt HB = square root-transformed sleep apnea-specific hypoxic burden.

In univariable Cox proportional hazards models (Table 2), sqrt HB, analyzed as either a continuous or categorical variable, was associated with the risk of the composite outcome (*p* = 0.043 and *p* = 0.002, respectively). In contrast, AHI, analyzed as a continuous variable, was not associated with the risk of the composite outcome, whereas AHI analyzed as a categorical variable was associated with the outcome (*p* = 0.066 and *p* = 0.013, respectively).

**Table 2.**
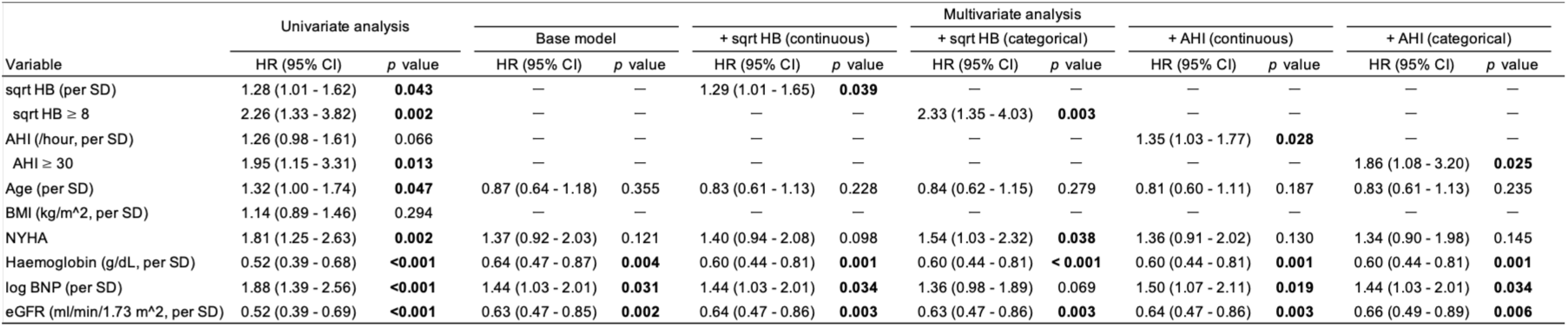
Univariable and Multivariable Cox Models for the Composite Outcome. Univariable and multivariable Cox proportional hazards models were used to evaluate associations with the composite outcome of cardiovascular death or rehospitalization for worsening HF. The baseline multivariable model included age, NYHA functional class, hemoglobin, log BNP, and eGFR, and models adding either sqrt HB or AHI to the baseline model were evaluated. All Cox models fulfilled the proportional hazards assumption. AHI = apnea-hypopnea index; BMI = body mass index; BNP = B-type natriuretic peptide; CI = confidence interval; eGFR = estimated glomerular filtration rate; HR = hazard ratio; NYHA = New York Heart Association; sqrt HB = square root-transformed sleep apnea-specific hypoxic burden.

After adjustment for 5 covariates—age as a potential confounder and NYHA functional class, hemoglobin, log BNP, and eGFR as clinically relevant factors (26,27)—sqrt HB and AHI remained independently associated with the composite outcome (Table 2). In these multivariable Cox proportional hazards models, for every 4.3-unit increase in sqrt HB, corresponding to 1 SD, the hazard of the composite outcome increased by 29% (HR = 1.29 [95% CI: 1.01–1.65]), corresponding to a 23.0 % increase in the estimated 5-year relative risk. For every 19.3-events/h increase in AHI, corresponding to 1 SD, the hazard of the composite outcome increased by 35% (HR = 1.35 [95% CI: 1.03–1.77]), corresponding to a 27.2 % increase in the estimated 5-year relative risk.

We further evaluated whether adding sqrt HB or AHI to the baseline multivariable Cox model improved prognostic performance. The baseline model included the 5 clinical covariates described above. The addition of sqrt HB improved model performance, as shown by an increase in the C statistic (bootstrap ΔC index: 0.020 [95% CI: 0.008 to 0.031]) and a decrease in the Brier score (bootstrap ΔBrier score: –0.007 [95% CI: –0.014 to –0.001]). In contrast, the addition of AHI did not clearly improve model performance (bootstrap ΔC index: 0.008 [95% CI: –0.003 to 0.016]; bootstrap ΔBrier score: –0.003 [95% CI: –0.009 to 0.001]) (Figure 3).

**Figure 3.**
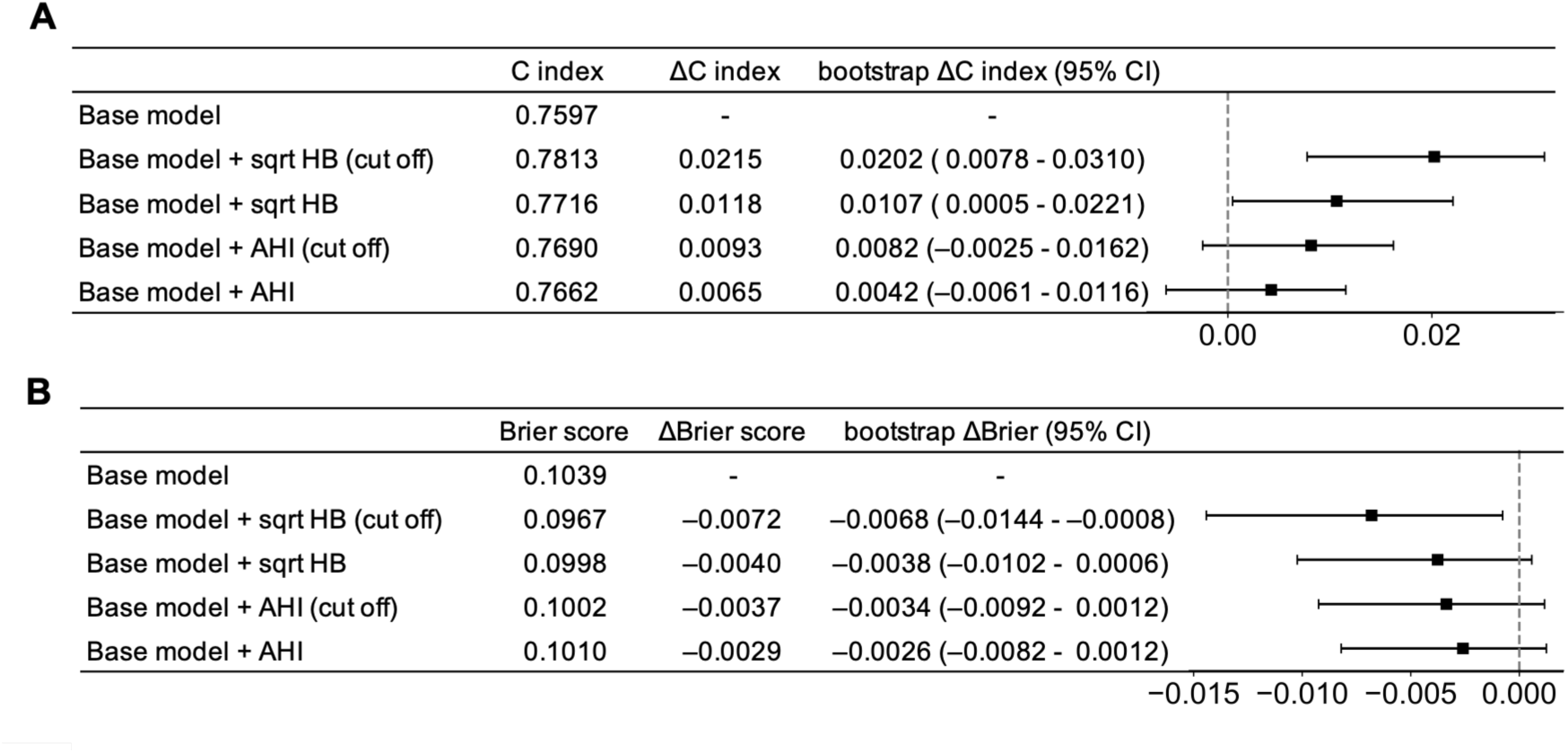
Prediction Improvement with Addition of sqrt HB and AHI. (A) C indices and (B) 5-year Brier scores are shown for the baseline model and models additionally including sqrt HB or AHI as cutoff-based or continuous variables. The baseline model included age, NYHA functional class, hemoglobin, log BNP, and eGFR. ΔC index and ΔBrier score indicate changes from the baseline model; higher C indices and lower Brier scores indicate better model performance. Brier scores were calculated using inverse probability of censoring weighting. Error bars indicate 95% CIs from 500 bootstrap resamples. AHI = apnea-hypopnea index; BNP = B-type natriuretic peptide; CI = confidence interval; eGFR = estimated glomerular filtration rate; NYHA = New York Heart Association; sqrt HB = square root-transformed sleep apnea-specific hypoxic burden.

In contrast, T90 ≥ 5 min was not associated with a higher cumulative incidence of the composite endpoint in Kaplan–Meier analysis (Supplemental Figure 3A). T90 was also not associated with the composite outcome when analyzed as either a continuous or categorical variable in univariable Cox proportional hazards models or after multivariable adjustment (Supplemental Table 2).

### Causal Inference Analysis Including PAP Treatment

Next, we analyzed the full cohort of 230 patients, including those who received PAP treatment. The baseline characteristics of the 230 patients are summarized in Supplemental Table 3. During follow-up, 9 patients (17.3%) in the PAP treatment group reached the composite endpoint. Kaplan–Meier curves showed lower event rates in patients treated with PAP, even among those with high sqrt HB or high AHI (sqrt HB ≥ 8 with vs without PAP treatment: *p* = 0.001; AHI ≥ 30 with vs without PAP treatment: *p* = 0.001) (Figure 4A, B). We then estimated the treatment effect using the g-formula (32). Among patients with sqrt HB < 8, the estimated 5-year risk was 29.4% without PAP and 27.4% with PAP, corresponding to an absolute risk reduction of 2.0%. Among those with sqrt HB ≥ 8, the corresponding risks were 45.8% and 26.6%, respectively, with an absolute risk reduction of 19.1%. Similarly, among patients with AHI < 30 events/h, the estimated 5-year risks were 30.6% without PAP and 30.8% with PAP, corresponding to an absolute risk reduction of −0.2%, whereas among those with AHI ≥ 30 events/h, the corresponding risks were 43.6% and 23.7%, with an absolute risk reduction of 19.9%. Consistent with these estimates, PAP-associated risk differences were close to zero in the lower sqrt HB and AHI categories but were more negative in the higher categories. The resulting interaction risk differences suggested that the association between PAP treatment and lower 5-year risk was greater in patients with higher sqrt HB or AHI (Figure 4C). In contrast, for T90, the PAP-associated risk difference was not statistically significant even in the higher category (≥ 5 min); the interaction risk difference was likewise not statistically significant (Supplemental Figure 3C).

**Figure 4.**
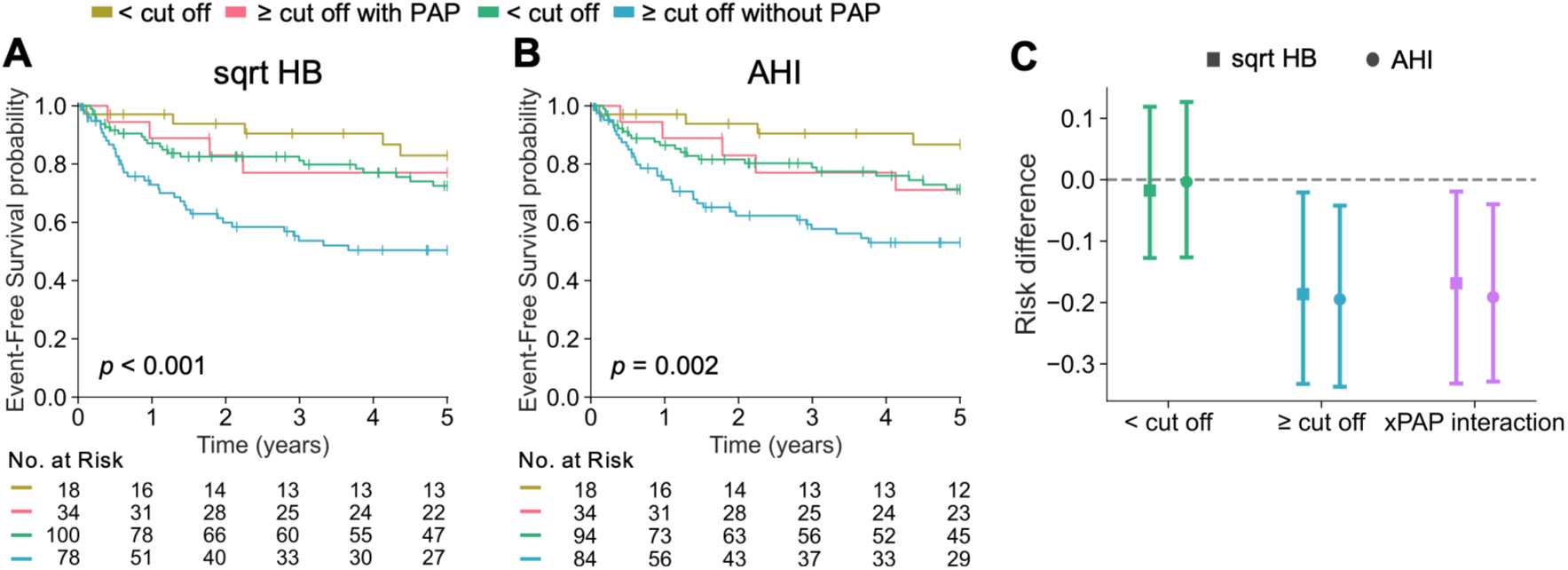
PAP Treatment Effects by sqrt HB and AHI. Kaplan–Meier event-free survival curves for the composite outcome of cardiovascular death or rehospitalization for worsening HF are shown according to PAP treatment and the cutoff categories of (A) sqrt HB 8 and (B) AHI 30 events/h. (C) PAP-associated 5-year risk differences and interaction risk differences estimated using the g-formula are shown. Negative values indicate lower risk with PAP treatment. Error bars indicate 95% CIs from 500 bootstrap resamples. AHI = apnea-hypopnea index; CI = confidence interval; sqrt HB = square root-transformed sleep apnea-specific hypoxic burden.

## Discussion

In this single-center observational cohort of 178 patients hospitalized for acute HF, SASHB was independently associated with clinical outcomes. The principal findings were as follows. First, although both SASHB and AHI were associated with the risk of cardiovascular death or rehospitalization for worsening HF, only SASHB improved model discrimination and prediction error when added to a model including established prognostic factors. Second, SASHB showed more consistent associations with the composite outcome than AHI across analyses. Third, in the analysis of 230 patients including those treated with PAP, both SASHB and AHI showed risk reduction associated with PAP treatment. These findings suggest that SASHB may provide additional prognostic information beyond AHI for risk stratification in hospitalized patients for HF.

The present findings extend prior observations in general populations and patients with coronary artery disease to patients hospitalized for HF. Previous studies evaluating SDB metrics in general populations showed that SASHB predicted cardiovascular mortality (14) and incident HF (15) more strongly than AHI. In patients with coronary artery disease, SASHB also predicted major adverse cardiac and cerebrovascular events better than AHI (16). These findings support the concept that event-related oxygen desaturation burden provides prognostic information not captured by event frequency alone. In the present study, SASHB was associated with adverse clinical outcomes in patients with HF, whereas AHI showed less consistent associations; patients with endpoint events had higher SASHB values, but no difference in AHI values, and continuous AHI was not associated with the composite outcome in univariable Cox models. SASHB improved risk stratification beyond established prognostic factors, whereas AHI did not improve model performance. These results suggest that the duration and depth of respiratory event-related desaturation may be particularly relevant for risk assessment in hospitalized patients with HF.

In addition, PAP treatment is commonly used to suppress SDB even in patients with HF and is known to be effective in a short-term (6). In the present study, both SASHB and AHI cutoffs showed interaction with PAP treatment, suggesting that patients with a greater SDB burden may derive greater risk reduction from PAP treatment. These findings suggest that SASHB, similar to AHI, may be useful for identifying HF patients who could benefit from PAP initiation. Because PAP treatment has not demonstrated a prognostic benefit in patients with moderate to severe SDB defined by AHI, a clinical trial enrolling patients with HF and high SASHB may be warranted.

The broader concept of hypoxic burden includes not only SASHB but also T90. Prior studies have shown that T90 is an independent predictor of all-cause mortality in HF (23–25). In the present study, however, T90 was not independently associated with the composite outcome and did not show risk reduction in the analysis of PAP treatment. One possible explanation is that T90 is not specific to individual respiratory events and may partly reflect underlying disease severity or baseline oxygenation status. In patients hospitalized for acute HF, individualized event-specific SASHB may therefore provide more clinically relevant prognostic information than T90.

Of note, this study evaluated SASHB in patients with HF, a population in which CSA is highly prevalent (36), and demonstrated that SASHB could be calculated for both OSA and CSA using the existing method. Patients with central-dominant SDB had a significantly longer desaturation duration than those with obstructive-dominant SDB, accompanied by a greater mean HB area per event. By incorporating the prolonged oxygen desaturation characteristic of central events, which is not fully captured by AHI alone, SASHB may provide additional prognostic information beyond AHI and thereby contribute to improved risk prediction in patients with HF.

## Study Limitations

This study has several limitations. First, because this was an observational study, unmeasured confounding may have influenced the outcomes even after multivariable adjustment. Second, although the g-formula was used for causal inference, the analysis should be interpreted cautiously because the number of patients with composite outcome events was very small in the PAP treatment group. Although interventions targeting both SASHB and AHI suggested potential outcome improvement, prior evidence in patients with coronary artery disease treated with PAP showed that SASHB predicted risk better than AHI (37), and the relative usefulness of SASHB and AHI for identifying patients who benefit from PAP treatment may therefore differ even in patients with HF, in larger studies with more outcome events. Third, because PAP treatment was not randomly assigned, selection bias cannot be excluded, and unmeasured differences between the groups or patients’ willingness to receive PAP treatment may have influenced the observed associations. Finally, because patients with HF with preserved ejection fraction were excluded, the present findings cannot be directly generalized to that population.

## Conclusions

In patients hospitalized for acute HF, SASHB showed a consistent and independent association with clinical outcomes. Although the prognostic utility of SASHB has been demonstrated primarily in populations with OSA, our findings suggest that it is also useful in patients hospitalized for HF, a population with a high prevalence of CSA. SASHB may provide additional prognostic information beyond AHI for risk stratification in this population.

## Supplementary Material

Supplemental Figures and Tables

## Abbreviations

AHI: apnea-hypopnea index
HF: heart failure
SDB: sleep-disordered breathing
SASHB: sleep apnea-specific hypoxic burden
PAP: positive airway pressure
NYHA: New York Heart Association
LVEF: left ventricular ejection fraction
CSA: central sleep apnea
OSA: obstructive sleep apnea
BNP: B-type natriuretic peptide

## References

1. Heidenreich PA, Bozkurt B, Aguilar D et al. 2022 AHA/ACC/HFSA Guideline for the Management of Heart Failure: A Report of the American College of Cardiology/American Heart Association Joint Committee on Clinical Practice Guidelines. J Am Coll Cardiol 2022;79:e263–e421.

2. Kitai T, Kohsaka S, Kato T, et al. JCS/JHFS 2025 Guideline on Diagnosis and Treatment of Heart Failure. Circ J 2025;89:1278–1444.

3. Khayat R, Jarjoura D, Porter K et al. Sleep disordered breathing and post-discharge mortality in patients with acute heart failure. Eur Heart J 2015;36:1463–9.

4. Kasai T, Kohno T, Shimizu W, et al. JCS 2023 Guideline on Diagnosis and Treatment of Sleep Disordered Breathing in Cardiovascular Disease. Circ J 2024;88:1865–1935.

5. Kober L, Adamo M, Ruwald AC et al. 2026 ESC Guidelines for the management of heart failure. Eur Heart J 2026.

6. Kato T, Suda S, Kasai T. Positive airway pressure therapy for heart failure. World J Cardiol 2014;6:1175–91.

7. Ishiwata S, Kasai T, Sato A et al. Prognostic effect of sleep-disordered breathing on hospitalized patients following acute heart failure. Clin Res Cardiol 2022;111:663–672.

8. Bradley TD, Logan AG, Kimoff RJ et al. Continuous positive airway pressure for central sleep apnea and heart failure. N Engl J Med 2005;353:2025–33.

9. Cowie MR, Woehrle H, Wegscheider K et al. Adaptive Servo-Ventilation for Central Sleep Apnea in Systolic Heart Failure. N Engl J Med 2015;373:1095–105.

10. O’Connor CM, Whellan DJ, Fiuzat M et al. Cardiovascular Outcomes With Minute Ventilation-Targeted Adaptive Servo-Ventilation Therapy in Heart Failure: The CAT-HF Trial. J Am Coll Cardiol 2017;69:1577–1587.

11. Bradley TD, Logan AG, Lorenzi Filho G et al. Adaptive servo-ventilation for sleep-disordered breathing in patients with heart failure with reduced ejection fraction (ADVENT-HF): a multicentre, multinational, parallel-group, open-label, phase 3 randomised controlled trial. Lancet Respir Med 2024;12:153–166.

12. Wang H, Parker JD, Newton GE et al. Influence of obstructive sleep apnea on mortality in patients with heart failure. J Am Coll Cardiol 2007;49:1625–1631.

13. Javaheri S, Shukla R, Zeigler H, Wexler L. Central sleep apnea, right ventricular dysfunction, and low diastolic blood pressure are predictors of mortality in systolic heart failure. J Am Coll Cardiol 2007;49:2028–34.

14. Azarbarzin A, Sands SA, Stone KL et al. The hypoxic burden of sleep apnoea predicts cardiovascular disease-related mortality: the Osteoporotic Fractures in Men Study and the Sleep Heart Health Study. Eur Heart J 2019;40:1149–1157.

15. Azarbarzin A, Sands SA, Taranto-Montemurro L et al. The Sleep Apnea-Specific Hypoxic Burden Predicts Incident Heart Failure. Chest 2020;158:739–750.

16. Peker Y, Celik Y, Zinchuk A, Sands SA, Redline S, Azarbarzin A. Association of Hypoxic Burden With Cardiovascular Events: A Risk Stratification Analysis of the Randomized Intervention With CPAP in Coronary Artery Disease and Sleep Apnea Cohort. Chest 2025;168:1481–1493.

17. McKee PA, Castelli WP, McNamara PM, Kannel WB. The natural history of congestive heart failure: the Framingham study. N Engl J Med 1971;285:1441–6.

18. Sato N, Kajimoto K, Asai K et al. Acute decompensated heart failure syndromes (ATTEND) registry. A prospective observational multicenter cohort study: rationale, design, and preliminary data. Am Heart J 2010;159:949–955 e1.

19. Berry RB AC, Harding SM, et al. The AASM Manual for the Scoring of Sleep and Associated Events: Rules, Terminology and Technical Specifications: American Academy of Sleep Medicine, Darien, IL, USA, 2018.

20. Matsuo S, Imai E, Horio M et al. Revised equations for estimated GFR from serum creatinine in Japan. Am J Kidney Dis 2009;53:982–92.

21. Lang RM, Badano LP, Mor-Avi V et al. Recommendations for cardiac chamber quantification by echocardiography in adults: an update from the American Society of Echocardiography and the European Association of Cardiovascular Imaging. J Am Soc Echocardiogr 2015;28:1–39 e14.

22. Kasai T, Motwani SS, Yumino D et al. Contrasting effects of lower body positive pressure on upper airways resistance and partial pressure of carbon dioxide in men with heart failure and obstructive or central sleep apnea. J Am Coll Cardiol 2013;61:1157–66.

23. Huang B, Huang Y, Zhai M et al. Association of hypoxic burden metrics with cardiovascular outcomes in heart failure and sleep-disordered breathing. ESC Heart Fail 2023;10:3504–3514.

24. Oldenburg O, Wellmann B, Buchholz A et al. Nocturnal hypoxaemia is associated with increased mortality in stable heart failure patients. Eur Heart J 2016;37:1695–703.

25. Pinna GD, Maestri R, Robbi E, Guazzotti G, Caporotondi A, La Rovere MT. Nocturnal hypoxemic burden in patients with heart failure: Emerging prognostic role of its nonspecific component. Am Heart J 2024;276:1–11.

26. Shiba N, Watanabe J, Shinozaki T et al. Analysis of chronic heart failure registry in the Tohoku district: third year follow-up. Circ J 2004;68:427–34.

27. Voors AA, Ouwerkerk W, Zannad F et al. Development and validation of multivariable models to predict mortality and hospitalization in patients with heart failure. Eur J Heart Fail 2017;19:627–634.

28. Harrell FE, Jr., Lee KL, Mark DB. Multivariable prognostic models: issues in developing models, evaluating assumptions and adequacy, and measuring and reducing errors. Stat Med 1996;15:361–87.

29. Graf E, Schmoor C, Sauerbrei W, Schumacher M. Assessment and comparison of prognostic classification schemes for survival data. Stat Med 1999;18:2529–45.

30. Gerds TA, Schumacher M. Consistent estimation of the expected Brier score in general survival models with right-censored event times. Biom J 2006;48:1029–40.

31. Steyerberg EW, Harrell FE, Jr., Borsboom GJ, Eijkemans MJ, Vergouwe Y, Habbema JD. Internal validation of predictive models: efficiency of some procedures for logistic regression analysis. J Clin Epidemiol 2001;54:774–81.

32. Denz R, Klaassen-Mielke R, Timmesfeld N. A comparison of different methods to adjust survival curves for confounders. Stat Med 2023;42:1461–1479.

33. Davidson-Pilon C. lifelines: survival analysis in Python. Journal of Open Source Software 2019;4:1317.

34. Virtanen P, Gommers R, Oliphant TE et al. SciPy 1.0: fundamental algorithms for scientific computing in Python. Nat Methods 2020;17:261–272.

35. Waskom ML. seaborn: statistical data visualization. Journal of Open Source Software 2021;6:3021.

36. Suda S, Kasai T, Matsumoto H et al. Prevalence and Clinical Correlates of Sleep-Disordered Breathing in Patients Hospitalized With Acute Decompensated Heart Failure. Can J Cardiol 2018;34:784–790.

37. Azarbarzin A, Vena D, Esmaeili N et al. Cardiovascular benefit of continuous positive airway pressure according to high-risk obstructive sleep apnoea: a multi-trial analysis. Eur Heart J 2026;47:2077–2089.

