## Supplemental Figures and Tables for "The Hypoxic Burden of Sleep Apnea Predicts Cardiovascular Outcomes in Patients with Heart Failure"

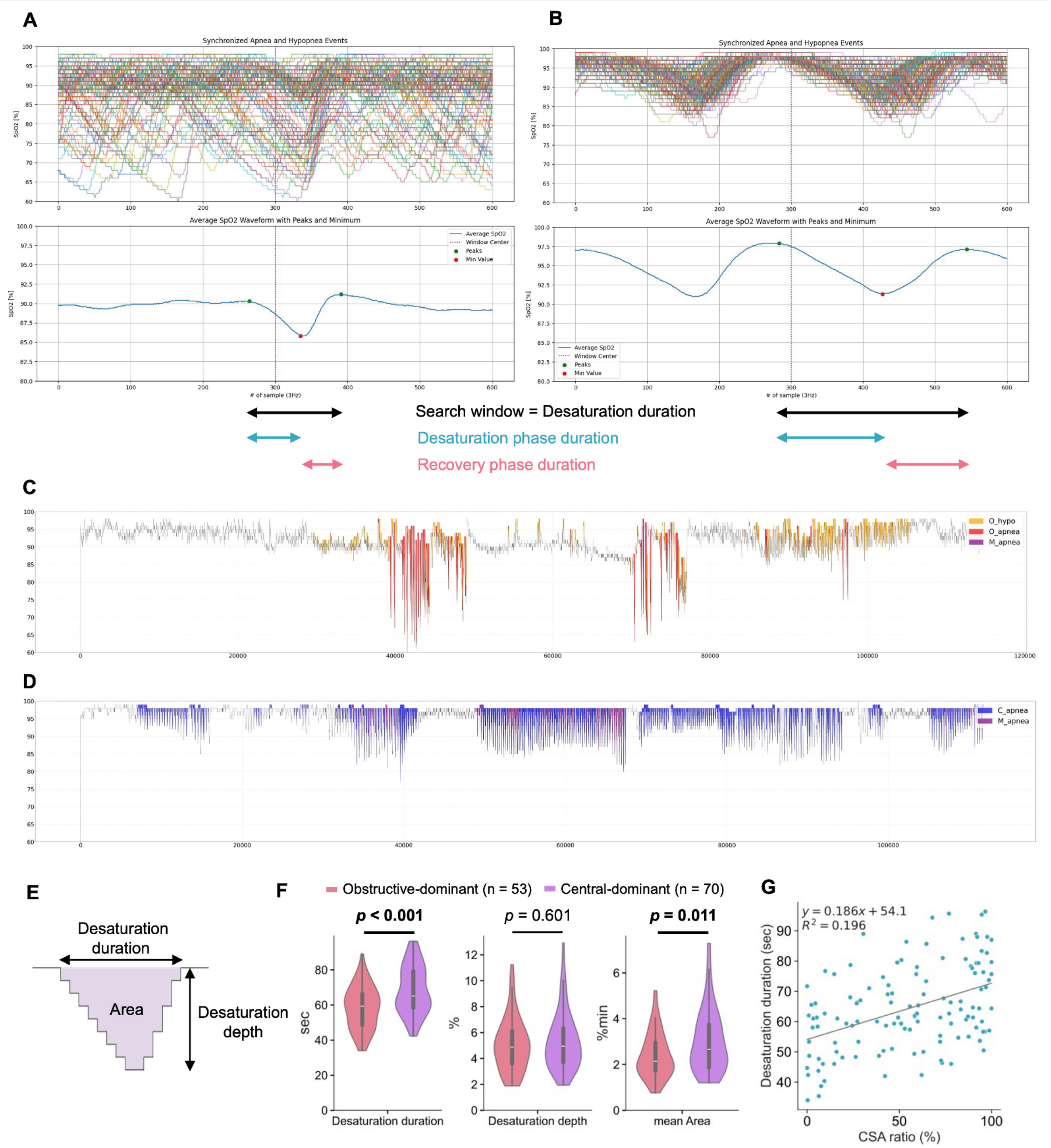


**Supplemental Figure 1. Hypoxic Burden Calculation and Desaturation Profiles in Obstructive-Dominant and Central-Dominant SDB**

(A, B) Representative $SpO_2$ segments from patients with (A) obstructive-dominant and (B) central-dominant SDB were aligned at time zero and averaged across the entire night to generate the mean desaturation curves. The patient in panel A had an AHI of 33.9 events/h, a SASHB of 134.5, and a CSA event proportion of 42.3%, whereas the patient in panel B had an AHI of 38.9 events/h, a SASHB of 161.0, and a CSA event proportion of 84.5%. Desaturation duration was divided into desaturation phase duration and recovery phase duration.

(C, D) Representative overnight $SpO_2$ recordings corresponding to panels A and B, respectively.

(E) Schematic illustration showing that mean area per event was approximated as desaturation duration × desaturation depth / 2.

(F) Comparison of desaturation duration, desaturation depth, and mean area per event between patients with obstructive-dominant and central-dominant SDB.

(G) Scatterplot showing the relationship between the proportion of CSA events and desaturation duration.

AHI = apnea-hypopnea index; CSA = central sleep apnea; SASHB = sleep apnea-specific hypoxic burden; SDB = sleep-disordered breathing; SpO2 = oxyhemoglobin saturation.

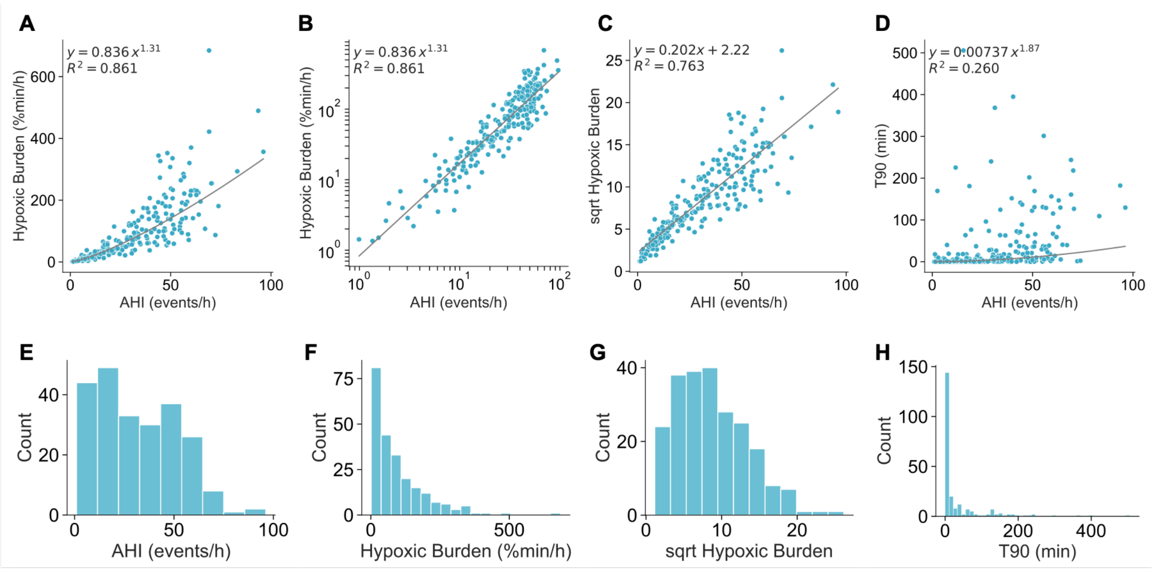


**Supplemental Figure 2. Scatter plots and histograms of SDB metrics**

(A–D) Scatterplots showing the relationships between AHI and (A) SASHB, (B) log-transformed SASHB, (C) sqrt HB, and (D) T90. In panel B, both SASHB and AHI were log-transformed.

(E–H) Histograms showing the distributions of the SDB metrics: (E) AHI, (F) SASHB, (G) sqrt HB, and (H) T90.

AHI = apnea-hypopnea index; SASHB = sleep apnea-specific hypoxic burden; sqrt HB = square root-transformed SASHB; T90 = time with oxygen saturation < 90%.

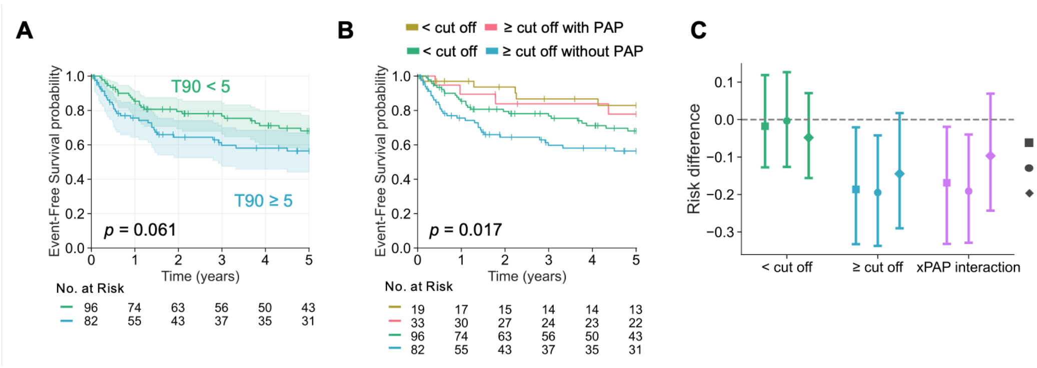


**Supplemental Figure 3. PAP Treatment Effects by T90 and Comparison of PAP-Associated Risk Differences**

(A) Kaplan–Meier event-free survival curves are shown according to the cutoff of T90 ≥ 5 min. Event-free survival for the composite of cardiovascular death or rehospitalization for worsening HF did not differ between patients with T90 < 5 min and those with T90 ≥ 5 min, as assessed using the log-rank test.

(B) Kaplan–Meier event-free survival curves for the composite outcome are shown according to PAP treatment and T90 categories of < 5 min and ≥ 5 min.

(C) PAP-associated 5-year risk differences and interaction risk differences estimated using the g-formula are shown for sqrt HB, AHI, and T90. Negative values indicate lower risk with PAP treatment. Error bars indicate 95% CIs from 500 bootstrap resamples.

AHI = apnea-hypopnea index; CI = confidence interval; SASHB = sleep apnea-specific hypoxic burden; sqrt HB = square root-transformed sleep apnea-specific hypoxic burden; T90 = time with oxygen saturation < 90%.

| | sqrt HB | | | AHI | | |
|---|---|---|---|---|---|---|
| | < 8 (n = 100) | ≥ 8 (n = 78) | *p* value | < 30 (n = 94) | ≥ 30 (n = 84) | *p* value |
| Patient characteristics | | | | | | |
| Age, year | 62.3 ± 13.4 | 65.9 ± 12.6 | 0.105 | 62.2 ± 13.3 | 65.7 ± 12.8 | 0.110 |
| Age ≥ 60, n(%) | 54 (54.0) | 57 (73.1) | **0.009** | 53 (56.4) | 58 (69.0) | 0.082 |
| Male sex, n(%) | 66 (66.0) | 61 (78.2) | 0.074 | 62 (66.0) | 65 (77.4) | 0.092 |
| BMI kg/m^2 | 22.6 ± 4.0 | 24.7 ± 5.2 | **0.011** | 22.5 ± 4.2 | 24.7 ± 4.9 | **0.002** |
| BMI ≥ 25 kg/m^2, n(%) | 23 (23.0) | 28 (35.9) | 0.059 | 21 (22.3) | 30 (35.7) | **0.049** |
| History of HF, n (%) | 48 (48.0) | 39 (50.0) | 0.791 | 46 (48.9) | 41 (48.8) | 0.987 |
| NYHA class at polysomnography | 2.3 ± 0.6 | 2.4 ± 0.6 | 0.176 | 2.3 ± 0.6 | 2.5 ± 0.6 | **0.031** |
| NYHA III/IV, n (%) | 26 (26.0) | 29 (37.2) | 0.109 | 22 (23.4) | 33 (39.3) | **0.022** |
| Systolic BP, mmHg | 104.0 ± 16.4 | 106.9 ± 15.9 | 0.177 | 102.9 ± 16.4 | 107.9 ± 15.7 | **0.038** |
| Diastolic BP, mmHg | 58.9 ± 10.4 | 61.1 ± 9.2 | 0.093 | 58.4 ± 10.3 | 61.5 ± 9.3 | **0.019** |
| LVEF, % | 34.6 ± 11.0 | 34.2 ± 10.1 | 0.904 | 34.1 ± 11.3 | 34.9 ± 9.7 | 0.319 |
| Diabetes, n (%) | 18 (18.0) | 32 (41.0) | **<0.001** | 17 (18.1) | 33 (39.3) | **0.002** |
| Composite outcome, n (%) | 23 (23.0) | 35 (44.9) | **0.002** | 23 (24.5) | 35 (41.7) | **0.015** |
| Laboratory variables | | | | | | |
| Haemoglobin, g/L | 13.5 ± 2.6 | 13.4 ± 2.2 | 0.992 | 13.5 ± 2.6 | 13.5 ± 2.3 | 0.730 |
| BNP, pg/ml | 357.7 ± 359.7 | 468.7 ± 404.4 | **0.015** | 373.4 ± 371.6 | 443.2 ± 394.0 | 0.124 |
| log BNP | 5.4 ± 1.0 | 5.8 ± 1.0 | **0.015** | 5.5 ± 1.0 | 5.7 ± 1.0 | 0.124 |
| eGFR, ml/min/1.73 m2 | 46.3 ± 22.0 | 41.9 ± 15.6 | 0.144 | 47.3 ± 21.7 | 41.1 ± 16.3 | **0.036** |
| eGFR < 60, n (%) | 73 (73.0) | 66 (84.6) | 0.063 | 68 (72.3) | 71 (84.5) | **0.050** |
| Sleep study finidings | | | | | | |
| Total recording time, min | 624.8 ± 72.3 | 620.8 ± 51.1 | 0.444 | 626.4 ± 70.2 | 619.3 ± 55.8 | 0.518 |
| Total sleep time, min | 361.0 ± 84.2 | 348.1 ± 83.9 | 0.346 | 365.0 ± 76.4 | 344.5 ± 91.2 | 0.186 |
| Sleep efficiency, % of TST | 58.2 ± 13.7 | 56.1 ± 12.8 | 0.325 | 58.7 ± 12.6 | 55.7 ± 14.1 | 0.275 |
| Wake after sleep offset, min | 136.2 ± 84.8 | 162.4 ± 72.8 | **0.006** | 132.4 ± 79.9 | 164.7 ± 78.4 | **0.002** |
| Slow wave sleep, min | 29.9 ± 28.7 | 20.1 ± 21.5 | **0.012** | 31.2 ± 28.5 | 19.4 ± 21.9 | **0.001** |
| REM sleep, min | 59.2 ± 30.3 | 50.4 ± 30.3 | **0.020** | 61.6 ± 28.6 | 48.4 ± 31.3 | **0.001** |

**Supplemental Table 1. Clinical Characteristics by sqrt HB and AHI Cutoffs**

Values are mean ± SD or n (%). Clinical characteristics were compared between patients below and above the cutoff values of sqrt HB 8 and AHI 30 events/h using the chi-squared test for categorical variables and the Mann–Whitney U test for continuous variables.

AHI = apnea-hypopnea index; BMI = body mass index; BNP = B-type natriuretic peptide; BP = blood pressure; eGFR = estimated glomerular filtration rate; HF = heart failure; LVEF = left ventricular ejection fraction; NYHA = New York Heart Association; REM = rapid eye movement; SD = standard deviation; sqrt HB = square root-transformed sleep apnea-specific hypoxic burden; TST = total sleep time.

| | Univariate analysis | | Multivariate analysis | | | | | |
|---|---|---|---|---|---|---|---|---|
| | | | Base model | | + T90 (continuous) | | + T90 (categorical) | |
| Variable | HR (95% CI) | *p* value | HR (95% CI) | *p* value | HR (95% CI) | *p* value | HR (95% CI) | *p* value |
| T90, min of TRT (per SD) | 1.08 (0.88 - 1.33) | 0.456 | — | — | 1.10 (0.88 - 1.38) | 0.385 | — | — |
| T90 ≥ 5 | 1.63 (0.97 - 2.74) | 0.063 | — | — | — | — | 1.37 (0.81 - 2.34) | 0.241 |
| Age (per SD) | 1.32 (1.00 - 1.74) | **0.047** | 0.87 (0.64 - 1.18) | 0.355 | 0.87 (0.64 - 1.18) | 0.367 | 0.85 (0.63 - 1.15) | 0.302 |
| NYHA | 1.81 (1.25 - 2.63) | **0.002** | 1.37 (0.92 - 2.03) | 0.121 | 1.21 (0.96 - 1.53) | 0.109 | 1.19 (0.94 - 1.50) | **0.145** |
| Haemoglobin (g/dL, per SD) | 0.52 (0.39 - 0.68) | **<0.001** | 0.64 (0.47 - 0.87) | **0.004** | 1.44 (1.03 - 2.01) | **0.033** | 1.42 (1.02 - 1.98) | **0.039** |
| log BNP (per SD) | 1.88 (1.39 - 2.56) | **<0.001** | 1.44 (1.03 - 2.01) | **0.031** | 0.64 (0.47 - 0.87) | **0.004** | 0.64 (0.47 - 0.86) | **0.004** |
| eGFR (ml/min/1.73 m^2, per SD) | 0.52 (0.39 - 0.69) | **<0.001** | 0.63 (0.47 - 0.85) | **0.002** | 0.63 (0.47 - 0.85) | **0.002** | 0.64 (0.48 - 0.86) | **0.003** |

**Supplemental Table 2. Univariable and Multivariable Cox Models for T90 and the Composite Outcome**

Univariable and multivariable Cox proportional hazards models were used to evaluate the association between T90 and the composite outcome of cardiovascular death or rehospitalization for worsening HF. The baseline multivariable model included age, NYHA functional class, hemoglobin, log BNP, and eGFR, and T90 was added to the baseline model as either a continuous or categorical variable using a cutoff of 5 min. All Cox models fulfilled the proportional hazards assumption.

BNP = B-type natriuretic peptide; CI = confidence interval; eGFR = estimated glomerular filtration rate; HR = hazard ratio; NYHA = New York Heart Association; T90 = time with oxygen saturation < 90%.

| Variable | Total (n = 230) | Event free (n = 163) | Incident case (n =67) | *p* value | Cohen's *d* |
|---|---|---|---|---|---|
| Patient characteristics | | | | | |
| Age, year | 62.8 ± 13.1 | 61.4 ± 13.1 | 66.1 ± 12.6 | **0.012** | 0.367 |
| Age ≥ 60, n(%) | 135 (58.7) | 86 (52.8) | 49 (73.1) | **0.004** | |
| Male sex, n(%) | 174 (75.7) | 127 (77.9) | 47 (70.1) | 0.213 | |
| BMI kg/m^2 | 24.1 ± 5.1 | 24.0 ± 5.0 | 24.3 ± 5.4 | 0.946 | 0.060 |
| BMI ≥ 25 kg/m2, n(%) | 78 (33.9) | 55 (33.7) | 23 (34.3) | 0.932 | |
| History of HF, n (%) | 113 (49.1) | 81 (49.7) | 32 (47.8) | 0.790 | |
| NYHA class at polysomnography | 2.3 ± 0.6 | 2.2 ± 0.5 | 2.5 ± 0.7 | **0.001** | 0.470 |
| NYHA III/IV, n (%) | 62 (27.0) | 34 (20.9) | 28 (41.8) | **0.001** | |
| Systolic BP, mmHg | 106.7 ± 16.5 | 106.2 ± 15.6 | 107.9 ± 18.5 | 0.896 | 0.103 |
| Diastolic BP, mmHg | 60.9 ± 10.4 | 60.9 ± 10.2 | 60.8 ± 11.0 | 0.902 | -0.012 |
| Heart rate, /min | 71.0 ± 12.5 | 70.8 ± 12.4 | 71.5 ± 12.6 | 0.825 | 0.056 |
| LVEF, % | 34.2 ± 10.6 | 34.5 ± 11.1 | 33.4 ± 9.4 | 0.589 | -0.101 |
| ICD, n (%) | 18 (7.8) | 8 (4.9) | 10 (14.9) | **0.010** | |
| CRT, n (%) | 16 (7.0) | 9 (5.5) | 7 (10.4) | 0.182 | |
| Comorbidities | | | | | |
| Atrial brillation, n (%) | 83 (36.1) | 54 (33.1) | 29 (43.3) | 0.145 | |
| Diabetes, n (%) | 67 (29.1) | 45 (27.6) | 22 (32.8) | 0.428 | |
| Laboratory variables | | | | | |
| Haemoglobin, g/L | 13.8 ± 2.5 | 14.3 ± 2.5 | 12.6 ± 2.1 | **< 0.001** | -0.708 |
| Sodium, mmol/l | 139.6 ± 3.2 | 139.7 ± 3.3 | 139.1 ± 3.0 | 0.076 | -0.209 |
| Potassium, mmol/l | 4.5 ± 2.5 | 4.6 ± 2.9 | 4.4 ± 0.5 | 0.309 | -0.068 |
| BNP, pg/ml | 387.8 ± 381.6 | 331.9 ± 346.1 | 523.6 ± 429.9 | **< 0.001** | 0.515 |
| log BNP | 5.5 ± 1.1 | 5.3 ± 1.1 | 5.9 ± 0.9 | **< 0.001** | 0.593 |
| eGFR, ml/min/1.73 m2 | 45.3 ± 19.1 | 48.8 ± 18.5 | 36.9 ± 18.0 | **< 0.001** | -0.652 |
| eGFR < 60, n (%) | 178 (77.4) | 119 (73.0) | 59 (88.1) | **0.013** | |
| Sleep study finidings | | | | | |
| Total recording time, min | 622.1 ± 60.9 | 624.2 ± 66.2 | 616.8 ± 45.5 | 0.563 | -0.122 |
| Total sleep time, min | 359.7 ± 82.6 | 371.2 ± 77.5 | 331.8 ± 88.5 | **0.002** | -0.488 |
| Sleep efficiency, % of TST | 58.0 ± 13.0 | 59.7 ± 12.1 | 54.0 ± 14.4 | **0.007** | -0.443 |
| Wake after sleep offset, min | 139.8 ± 80.3 | 126.8 ± 72.5 | 171.6 ± 89.5 | **< 0.001** | 0.576 |
| Slow wave sleep, min | 25.3 ± 25.5 | 28.2 ± 27.1 | 18.3 ± 19.4 | **0.011** | -0.394 |
| REM sleep, min | 57.4 ± 30.7 | 61.1 ± 31.1 | 48.4 ± 27.9 | **0.002** | -0.423 |
| AHI, /h of sleep | 31.7 ± 20.0 | 30.8 ± 20.0 | 33.9 ± 20.1 | 0.278 | 0.153 |
| HB, %min/h of sleep | 96.2 ± 99.1 | 92.3 ± 98.1 | 105.7 ± 101.8 | 0.242 | 0.135 |
| sqrt HB | 8.6 ± 4.6 | 8.4 ± 4.6 | 9.2 ± 4.7 | 0.242 | 0.154 |
| 3% ODI, /h of sleep | 24.6 ± 18.6 | 23.7 ± 18.2 | 26.9 ± 19.5 | 0.304 | 0.175 |
| T90, min of TRT | 35.1 ± 70.5 | 34.7 ± 74.2 | 36.2 ± 60.9 | 0.383 | 0.020 |
| Obstructive-dominant, n (%) | 77 (33.5) | 54 (33.1) | 23 (34.3) | 0.861 | |
| Medication at discharge | | | | | |
| ACE-Is/ARBs | 188 (81.7) | 132 (81.0) | 56 (83.6) | 0.643 | |
| Beta blocker | 214 (93.0) | 149 (91.4) | 65 (97.0) | 0.129 | |
| Aldosterone antagonist | 146 (63.5) | 105 (64.4) | 41 (61.2) | 0.645 | |
| Loop diuretics | 186 (80.9) | 125 (76.7) | 61 (91.0) | **0.012** | |

**Supplemental Table 3. Clinical Characteristics by Endpoint Event Status**

Values are mean ± SD or n (%). Clinical characteristics were compared between patients with and without endpoint events of cardiovascular death or rehospitalization for worsening HF, using the chi-squared test for categorical variables and the Mann–Whitney U test for continuous variables.

ACE-I = angiotensin-converting enzyme inhibitor; AHI = apnea-hypopnea index; ARB = angiotensin II receptor blocker; BMI = body mass index; BNP = B-type natriuretic peptide; BP = blood pressure; CRT = cardiac resynchronization therapy; eGFR = estimated glomerular filtration rate; HF = heart failure; ICD = implantable cardioverter defibrillator; LVEF = left ventricular ejection fraction; NYHA = New York Heart Association; ODI = oxygen desaturation index; REM = rapid eye movement; SASHB = sleep apnea-specific hypoxic burden; SD = standard deviation; sqrt HB = square root-transformed SASHB; T90 = time with oxygen saturation < 90%; TRT = total recording time; TST = total sleep time.
